# Assessment of Potentially Toxic Trace Elements in commonly consumed fruits and vegetables from Supermarkets and Associated Health Risks in Thulamela Municipality, South Africa: A study protocol

**DOI:** 10.64898/2026.10.26.26366572

**Authors:** Ndivhuwo Eunice Bulwana, Ngoako Solomon Mabapa

**Affiliations:** Department of Nutrition, Faculty of Health Sciences, University of Venda, Thohoyandou, South Africa

**Keywords:** Cadmium, fruits and vegetables, ICP-MS, Health risk assessment, Thulamela Municipality

## Abstract

Potentially toxic trace elements (PTTEs) contamination is a major global food health burden, with lead exposure causing more than 33 million disability-adjusted life years lost in 2021, mainly from cardiovascular effects. Cadmium in food is strongly linked to kidney dysfunction and reproductive toxicity, while arsenic and zinc also pose serious health risks. Exposure occurs through industrial emissions, contaminated air and water. Despite global studies, local data on these elements in South African fruits and vegetables remain limited. The study aims to determine the levels of arsenic, lead, cadmium, and zinc in fruits and vegetables from Thulamela Municipality and assess potential health risks to consumers and compare the levels of the elements to legal requirements.

A cross-sectional design will be adopted. Ten commonly consumed fruits and vegetables will be sampled purposively from three supermarkets in Thulamela and analysed for PTTEs using Inductively Coupled Plasma Spectrometry (ICP-MS) at University of Venda, and assessment of dietary exposure among adult consumers aged 18-65 years through 24-hour recall and Quantified Food Frequency Questionnaire (QFFQ). Differences in PTTEs with metal type and food type as factors will be determined using two-way ANOVA. Dietary data will be coded and analysed using Food Finder 3 software linked to SAFOODS to estimate intake. Health risks associated with intake of the PTTEs will be evaluated in terms of estimated daily intake (EDI), target hazard quotient (THQ), hazard index (HI) and Carcinogenic Risk (CR). Statistical analysis will be done using IBM SPSS version 29, and significance set at p<0.05. Measured elements levels will be compared to national legal limits to estimate daily intake and evaluate health risks. Ethical approval will be obtained and participants’ rights to consent, privacy, and confidentiality, will be protected.

Expected outcomes include contamination levels of PTTEs in selected fruits and vegetables and associated health risk assessment.

## Introduction and background

Adequate consumption of vegetables and fruits can save the lives of an estimated number of 2.7 million people. Over the years, there has been an increased awareness creation on the health benefits of vegetables. This has led to a substantial in vegetable consumption across the globe. They are consumed in both cooked and raw forms, thus vegetables containing potentially toxic trace elements (PTTEs) can cause detrimental effects on human health [1]. For good health, vegetables are supposed to be safe for consumption. Food safety is a major public health concern globally. Contamination with PTTEs is very important, particularly in agricultural production systems. There has been considerable attention paid to the issue of environmental pollution as one of the most critical global concerns. In recent decades, there has been noticeable degradation of the quality of air, water, and soil resources in different countries because of human activities. Therefore, several contaminants have been introduced into food chains, including pesticides and PTTEs [2]. However, the quality of vegetables consumed has been compromised, particularly in developing countries. This poses a crucial health risk to consumers, particularly because most vegetables are eaten raw. Parameters affecting the concentration of PTTEs in crops include climate, environmental pollution, type of soil, and the maturity degree of the plant at the time of harvesting [3]. One of the major and indirect pathways through which PTTEs enter the human food chain is through the absorption of PTTEs by plants growing on contaminated agricultural lands [4]. Accumulation of PTTEs such as Cadmium (Cd), nickel (Ni), and lead (Pb) in fruits and vegetables is one of the most important environmental issues that received much attention. Various reports have indicated the occurrence of PTTEs in vegetables in many countries including Pakistan, India and Ethiopia [5]. Consumption of PTTEs through the food chain has been thoroughly documented worldwide [6]. Monitoring levels of PTTEs can provide useful information for promoting food safety in South African food industries. In the Limpopo Province, Moyo et al found that soils and vegetables in Thohoyandou, located in Thulamela Municipality, are contaminated, posing a potential risk to consumers purchasing from local markets. Nevertheless, there is still a lack of data regarding PTTEs in fruits and vegetables available in formal supermarkets within Thulamela. Contradictory results, however, might be explained by the methodological differences among the South African research that are now available. Using a limited sample from informal markets using graphite furnace AAS, Moyo et al. detected Cd values of 0.23-2.94 mg/kg in vegetables from Thohoyandou without supermarket surveillance or health risk assessment.[7] On the other hand, according to FAO/WHO criteria, the study conducted in household gardens in Alice found that Pb was undetectable and Cd was 0.01-1.12 mg/kg using ICP-OES and health risk indices (THQ, HI) is justified by these differences in analytical technique (AAS vs. ICP-OES), sampling site (home gardens vs. informal markets), and lack of formal supermarket data.[8] There are several trace elements that are well known for being toxic, non-biodegradable, and potentially bio-accumulative in human tissues [9]. Several studies have demonstrated that excessive PTTEs intake can result in organ damage and cause multiple diseases, such as osteomalacia, renal and nerve dysfunction, and cardiovascular problems [10]. Diet is the main exposure route to PTTEs for humans. Therefore, the tolerate daily intake of these elements in food was considerably restricted by authorities and organizations [11]. Among them, Cd and Pb are highly toxic trace elements that, even at very low levels, can cause kidney damage and may lead to reduced reproductive ability, liver dysfunction, high blood pressure, and tumours [12]. Lead (Pb) exposure causes a significant burden of disease. The Institute for Health Metrics and Evaluation (IHME) estimates more than 1.5 million deaths globally were attributed to lead exposure in 2021, primarily due to cardiovascular effects. Additionally, lead exposure was estimated to account for more than 33 million years lost to disability (disability-adjusted life years, or DALYs) worldwide in 2021 [13]. Trivalent and inorganic compounds of arsenic (As) are more hazardous than organic, pentavalent compounds. Arsenic is a carcinogen that has a variety of acute and long-term detrimental impacts on human health [14]. Ingestion, inhalation, and skin absorption are some of the critical routes for arsenic entering human body. Both pentavalent and trivalent arsenic compounds are extensively and rapidly absorbed from the gastrointestinal tract. Several health problems are related to arsenic exposure, ranging from the development of cancer, inducing epigenetic changes, to skin diseases [15]. Long-term cadmium (Cd) exposure may cause various adverse effects involving kidney tubular disorders, resulting in the dysfunction of channels and transporters of the renal tubular system, resulting in the loss of fluids and a shift in electrolyte homeostasis [16]. Nowadays it is also known that zinc (Zn), especially zinc deficiency, plays an important role in many metabolic and chronic diseases, including diabetes, cancer (e.g., oesophageal, hepatocellular, breast cancer, colon cancer), neurodegenerative diseases such as Alzheimer’s disease, and intestinal diseases such as inflammatory bowel disease and irritable bowel syndrome [17]. In this light, it is also important to note that excessive intake of this element can have negative effects, such as disturbances in the levels of other elements, particularly copper, which can subsequently result in copper deficiency anaemia, reduction in the copper-dependent enzymes and cholesterol metabolism dysregulation [18]. Zinc will be included in the study because excessive dietary exposure may pose adverse health effects despite its physiological role as an essential trace element. Health evaluators such as the National Institute of Health (NIH) directed daily consumption of 8 mg Zn for adult females and 11 mg for adult males [19]. To protect consumer health, the FAO/WHO Codex Alimentarius Commission has set maximum levels (MLs) for PTTEs in vegetables and fruits such as inorganic arsenic (As) 0.2 mg/kg in rice(polished), cadmium (Cd) 0.20 mg/kg in leafy vegetables, lead (Pb) 0.30 mg/kg in leafy vegetables and 0.1 mg/kg in fruits with zinc (Zn) not regulated [20]. Therefore, the WHO/FAO permissible limit of 99.4 mg/kg for zinc as reported in recent studies will be adopted in this study [21].

### Problem Statement

The consumption of fruits and vegetables contaminated by PTTEs can deplete essential nutrients in the body and has been recognized as a risk to human health [22]. Ingestion of these elements can bring about weakening of the immune system, psycho-social dysfunctions, gastro-intestinal cancer, cardiovascular, renal, neurological and bone disease [23]. Globally, PTTE contamination of staple food is a documented crisis. A 2025 study by Health Babies Bright Futures tested 145 rice products across the United States and found 100% contained arsenic (As), while cadmium (Cd) was detected in all but one sample.[24]

Similar contamination in home garden systems has been reported in South Africa. Vegetables grown in home gardens in Alice, Eastern Cape Province, South Africa, were found to be contaminated with PTTEs. Manganese concentrations were extremely high in spinach (112.38 mg/kg) and soils (377.61-499.68 mg/kg), while zinc concentrations in spinach, onions, and tomatoes exceeded safe limits. The study showed that home grown vegetables in South Africa can acquire hazardous metals from local soils, even when lead and cadmium levels were below FAO/WHO guidelines [8]. In Limpopo Province, Thulamela Municipality within Vhembe District exhibits comparable agricultural practices, where smallholder farmers utilize locally produced compost manure, kraal manure, and sewage sludge from Thohoyandou Wastewater Treatment plant (WWTP). [25] Previous studies have identified the presence of Cd, Mn, and Ni in fruits and vegetables from Thohoyandou town, resulting in a knowledge deficiency that obstructs initiatives aimed at ensuring food safety and safeguarding public health in Thulamela.[7][8]

While strict regulations exist globally to monitor PTTEs, environmental and industrial pollution are recognised as primary entry routes into the food chain [26]. Thulamela lacks sufficient localized surveillance data to determine whether these regulations are effective or if exposure is occurring. Without continuous monitoring and research into agricultural practices, the extent of contamination remains unknown, leaving vulnerable populations unable to make informed dietary choices regarding produce purchased from supermarkets in Thulamela.

### Significance of the study

This study will contribute to the limited body of knowledge on PTTEs contamination in food crops in South Africa, specifically in the Thulamela municipality. Currently, few studies have linked laboratory analysis of PTTEs in produce to actual dietary intake in the area. While limited studies have indicated that concentrations of PTTEs in fruits, vegetables, and soils in the region can exceed recommended maximum acceptable levels, posing a health risk to consumers including potential kidney, neurological, and cancer risks from chronic exposure [7]. By assessing the contamination of these elements, the findings will inform policy makers and regulatory bodies such as Thulamela Environmental Health Department and Department of Agriculture on the safety of locally produced fruits and vegetables in Thulamela municipality and raising awareness among farmers, vendors, and consumers about health risks, and guide future research on food safety and PTTE contamination.

### Purpose of the study

To determine the concentrations of arsenic, cadmium, lead, and zinc in commonly consumed fruits and vegetables obtained from supermarkets in Thulamela Municipality, compare detected concentrations with national and international permissible limits, and assess the potential health risks associated with their consumption.

### Objectives of the study

a. To quantify concentrations of arsenic (As), cadmium (Cd), lead (Pb), and zinc (Zn) in selected fruits and vegetables obtained from supermarkets in Thulamela Municipality.
b. To compare measured concentrations with WHO/FAO Codex Alimentarius and South African regulatory limits.
c. To estimate dietary exposure to PTTEs among adults residing in Thulamela Municipality.
d. To assess non-carcinogenic and carcinogenic health risks using EDI, THQ, HI, and TCR models.

### Research Questions

a. What are the concentrations of As, Cd, Pb and Zn in selected fruits and vegetables sold in supermarkets in Thulamela Municipality?
b. Do measured concentrations exceed internationally accepted permissible limits?
c. What is the estimated dietary exposure to these PTTEs among adults in Thulamela Municipality?
d. What non-carcinogenic and carcinogenic health risks are associated with consumption?

### Hypothesis

#### Null Hypothesis

H₀: Concentrations of As, Cd, Pb and Zn in fruits and vegetables do not exceed permissible limits and do not pose significant health risks.

#### Alternative Hypothesis

H₁: Concentrations of As, Cd, Pb and Zn in fruits and vegetables exceed permissible limits and may pose significant health risks to consumers.

## Materials and Methods

### Study Design

The study will adopt a cross-sectional analytical design. This design will employ quantitative chemical analysis and dietary assessment methods. The design will allow for the determination of PTTEs concentrations in fruits and vegetables from Thulamela municipality markets using Shimadzu ICP-MS 2040, and the assessment of dietary exposure among adult consumers through a 24-hour recall and a Quantified Food Frequency Questionnaire. The study will also focus on PTTEs and compare them to legal limits. Dietary data will also be used to figure out how much people eat each day; how dangerous it is and how likely it is to cause cancer and to evaluate the potential health implications for the local population.

### Study Setting

The study will focus on Thulamela municipality within Vhembe District, a district in the Limpopo Province of South Africa that is farthest north, sharing a border with Botswana and Zimbabwe in the north-east and Mozambique in the south-east through parts of the Kruger National Park [27]. It is the smallest municipality of the four municipalities in the Vhembe district in terms of its geographical area, making up only 10% of the district. However, it is the largest municipality in the province in terms of population. The name Thulamela is a Karanga word meaning “the place of giving birth’’. The area is 2642 km ² and according to Statistics South Africa Census, Thulamela has a total population of 575,929, of which 355,433 (61,7%) are adults aged 15-64 years who represents the primary consumer age group for fruits and vegetables bought in supermarkets [28].

The Vhembe District forms a significant component of the production of several subtropical crops that contribute to the country’s agricultural economy particularly through exports. The well-established white commercial horticulture farming is generally found in the Southern side of the district [29]. It is mainly made up of stakeholders in the subtropical industry which includes commodities such as litchis, bananas, mangos, avocados, citrus, and pecan nuts. The district’s commercial horticulture farming, especially in the southern side, makes it an important area for assessing PTTE (arsenic (As), lead (Pb), cadmium (Cd) and zinc (Zn)) contamination in fruits and vegetables due to potential risks from agricultural practices and environmental pollution.

### Study population and sampling

#### Food Sample Collection

Purposive sampling techniques will be used to select three supermarkets in the Thulamela Municipality based on their high customer traffic and the consistent availability of the targeted fruits and vegetables. From each supermarket 10 types of fresh fruits and vegetables commonly consumed in the area will be purchased. For each commodity, 3 batches will be collected per supermarket on 3 separate occasions, with 1kg per batch, resulting in a total of 90 samples (3 supermarkets ×10 commodities ×3 batches). Each batch will be analysed in triplicate to ensure reliability. No pooling or composite sampling will be performed.

### Human participants

#### Sample Size Determination

The sample size was determined using the prevalence-based sample size calculation formula. An expected prevalence of 33% was adopted from a study conducted which reported fruit and vegetable consumption practices among rural communities in Limpopo province [30].

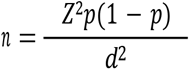

Where:

- n = required sample size132ewdxzc
- Z = Z-score corresponding to the desired confidence level (1.96 for 95% confidence)
- p = expected prevalence
- d = margin of error (precision)

Substituting the values into the formula:

- Expected prevalence = 33% (p = 0.33)
- Confidence level = 95% (Z = 1.96)
- Margin of error = 5% (d = 0.05)

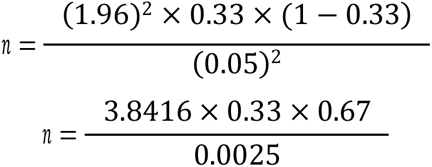

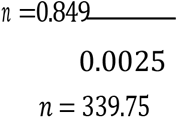

The calculated minimum sample size was approximately 340 participants. To compensate for potential non-response and attrition, an additional 10% was added:

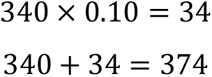

Therefore, the final sample size was rounded to 374 participants.

### Sampling technique

Convenience sampling will be used at selected supermarkets premises. Participants will be recruited at the exit points after they are done with shopping. They will be approached during data collection periods, which will encompass weekdays and weekends (09:00 to 17:00), potential participants one after the other will be approached to capture various groups of customers. The aim of the study will be explained to each prospective participant, and before registration, those who fit the inclusion requirements and are willing to take part will be asked to give written informed consent.

### Inclusion Criteria

Adults aged 18-65 residing in Thulamela Municipality, Fresh fruits and vegetables commonly consumed and sold within Thulamela Municipality supermarkets and Participants able to communicate in English and Tshivenda willing to take part in the study will be recruited

### Exclusion Criteria

Individuals under 18 years or over 65 years and those who will refuse to participate will be excluded

### Sample Preparation

The samples of fruits and vegetables will be collected from supermarkets in Thulamela municipality within Vhembe District and brought to the laboratory of nutrition at University of Venda, for the sample’s preparations. The common names and scientific names of the samples are Pyrus (pears), Musa acuminata (bananas), Solanum Lycopersicum (tomatoes), Malus domestica (apples), Vitis vinifera L (grapes) as well as Brassica oleracea(cabbage), Spinacia oleracea (spinach), Cucurbita moschata (thanga), black night shade (muxe), Brassica juncea (mutshaini), these commodities were selected because they are among the most commonly consumed fruits and vegetables in Thulamela municipality.

The fruits and vegetables collected will be rinsed with distilled water properly to remove the contaminated particles. After that they will be chopped to make nearly uniform small pieces. Then, these small pieces of fruits and vegetables will be placed in the acid washed clean and labelled proclaim crucibles. Later, the small pieces of fruits and vegetables will be dried in oven at 50°C for several hours to remove moisture until they become stiff and crispy.

The dried Vegetables and fruits pieces will be crushed and grounded by using mortar and piston to fine powder to get homogenized samples. Later, the dried powdered samples will be stored in polyethylene bags for further process.

### Digestion using microwave digestion system

Digestion: Approximately 1.0 g of accurately weighed dry ground samples will be taken into a Polytetrafluoroethylene (PTFE) microwave digestion vessel. 10 ml of concentrated nitric acid (HNO_3,_ 65%) and 2ml of hydrogen peroxide (H_2_O_2,_ 30%) will be added to each vessel. The vessels will be sealed and digested in a microwave digestion system with a ramp to 180°C over 15 minutes held, then cooled to room temperature. The digested samples will be filtered into 50 ml volumetric flask through Whatman filter paper no 42 and will be diluted with de-ionized water up to the mark of 50ml labelled the flask and kept at 4°C for further analysis of potentially toxic trace elements in fruits and vegetables using ICP-MS.

### ICP-MS instrumental parameters of analysis

Analytical measurements of the elements (Zn^65^, As^75^, Cd^111^, Pb^208^) will be performed using a Shimadzu ICPMS 2040) (iCAP Q ICP-MS Thermo Fisher Scientific, Waltham, Massachusetts, USA), a technique for detecting metals and several non-metals at very low concentrations. The ICP-MS is equipped with an ASX-520 autosampler, a micro-concentric nebulizer, cones (the instrument uses Ni sampler and Ni skimmer cones), and a peristaltic sampled delivery pump, and running a quantitative analysis mode. A nebulizer fitted to a cyclonic spray chamber will be used for introducing the sample solutions into ICP-MS plasma; the standard ICP-MS torch includes a 1.5 mm diameter injector.

Before the quantitative evaluation of the sample, the ICP-MS will be allowed to equilibrate (from the star up) for at least 45 min; during this time the experimental conditions and mass calibration of the instrument will be checked. A short-time stability test will be performed using a tuning standard solution (TUNE B iCAP QBa, Bi, Ce, Co, In, Li, U–1.0 µg/L (each) in 2% HNO_3_ + 0.5 HCl)) covering the whole range of masses. The role of this autotune is to allow optimizations of the sampling zone of the plasma to provide a compromise between high sensitivity, optimizations of ions optics voltage, and low levels of cluster ions and doubly charged ions. The ICP-MS will be optimized daily to give a maximum sensitivity for M+ ions, and the double ionization and oxides will be monitored by the means of the ratio between Ba^2+^/ Ba^+^ and Ce^2+^/ CeO^+^, respectively; these will always be less than 2%. The argon (Ar 5.0) and helium (He 6.0) used will be 99.99% purity (Messer, Austria). Each sample will be analysed in duplicate, and each analysis will consist of seven replicates.

### Reagents and Solutions

All reagents used will be analytical grades: 65% HNO_3_–supra pure for trace analysis (Merk, Darmstadt, Germany), and H_2_O_2_ ≥ 30% for trace analysis (Sigma-Aldrich, Stein Heim, Germany). The analysis will be made using the external standard calibration method, after an appropriate dilution and ICP-MS technique [31]. Each sample will be analysed in duplicate, and each analysis will be prepared in three replicates [32]. The standard solution used for generating the calibration curve and for the calibration of ICP-MS was high purity ICP Multi-Element Standard Solution XXI CertiPUR (Merk, Darmstadt, Germany). Ge, Tb, Rh, and Sc in supra pure 1% HNO3 (Merk, Darmstadt, Germany) were used as internal standards (the internal standard was added to the level of 50 µg/L to all samples, blanks, and standards). For microwave digestion, a mixture of concentrated nitric acid (HNO_3_, 65%) and hydrogen peroxide (H_2_O_2_, 30%) will be employed.

### Quality Control

Appropriate quality assurance procedures and precautions will be taken to ensure the reliability of the results. The glassware used for the storage or processing of the samples will be properly cleaned, and the handling of the samples will be performed carefully to avoid cross-contamination. All glassware and plasticware/polyethylene bottles used for sample preparation and storage will be cleaned by soaking in 10% v/v HNO3 for 24 h and rinsing at least three times with distilled water. The Teflon vessel used for sample digestion will also be cleaned by soaking in 5% HNO3 for more than 24 h, then rinsed with distilled water and dried at 105 ◦C in the oven. To implement the quality control of the chemical analyses, the blank method will be used through the complete preparation and analytical procedure. The blank spike duplicate (BSD) will be performed in triplicate to confirm the procedure is working within the established control limits. Control samples and working standards will be prepared daily from the intermediate standards (the intermediate solutions stored in polyethylene bottles will be prepared from the stock solution) [33].

Before starting the PTTE analysis sequence, mass calibration will be performed using a 250 mL setup solution containing Pb, Zn, Cd and As (3–35 µg/L in 2%HNO3), and the relative standard deviation (RSD < 5%) will be checked using a tuning solution 500 mL (TUNE B I CAP Q Ba, Bi, Ce, Co, In, Li, U—1.0 µg/L (each) in 2%HNO3 +0.5HCl) purchased from Thermo Scientific. The certified reference materials (CRM) will be used to check the accuracy of the analyses, the methodology used for verifying the accuracy consisted of replicating analyses of known sample concentrations of CRMs, and the obtained values ranged between 0.1% and 4.0%, depending on the element, the global recovery for each element was between 97% and 102%.

To establish the quality control of the analyses performed, blanks and triplicates samples (n = 3) will be analysed during the procedure, and the variation coefficient will be kept under 5%. One calibration blank (1% HNO3) will be used to establish the analytical curve, and detection limits (ppb) will be determined by the calibration curve method. Method blanks will be used to check for possible sample contamination during the sample’s preparation procedure. For each analysed sample, the spike method will be performed by analysing the control and the standard samples. Limit of detection (LoD) and limit of quantification (LoQ) will be calculated using the following mathematical equations: LoD = 3 × SD/s and LoQ=10×SD/s, where SD=estimation of the standard deviation of the regression lines, and s=slope of the calibration curve).

The calibration standard solution will be prepared from multi-element standard solution ICP Multi-Element Standard Solution XXI CertiPUR (obtained from Merk, Darmstadt, Germany), in seven concentrations ranging between 2.5 µL, 5 µL, 10 µL, 25 µL, 50 µL, 100 µL, and 250 µL. As presented, the accuracy of the analytical procedure will be investigated by spiking a suitable known amount of the analyte metal into a test portion of the sample and analysing the spiked test portion along with the original samples [34], while the precision of the analytical procedure will be expressed as RSD% of the triplicate analyses [35].

### Data Collection instrument

Two dietary assessment instruments will be used to collect dietary data. The first instrument, a 24-Hour Dietary Recall (24HR), will be used to identify the fruits and vegetables most commonly consumed by participants in Thulamela Municipality, including those purchased from supermarkets. Information obtained from the 24HR will be used to develop a list of commonly consumed fruits and vegetables, which will subsequently be incorporated into the Quantified Food Frequency Questionnaire (QFFQ).

The second instrument, a Quantified Food Frequency Questionnaire (QFFQ), will be used to assess participants’ usual fruit and vegetable consumption patterns over the preceding month. The QFFQ will consist of a finite list of fruits and vegetables commonly consumed in Thulamela Municipality, identified through the 24HR, and will include closed-ended questions to quantify the frequency and amount of consumption.

### Dietary Exposure Assessment

- To capture day-to-day variation in dietary patterns, the 24HR will be administered three times per participant on non-consecutive days, including at least one weekend day and two weekdays. Interviews will be conducted face-to-face using a multiple-pass technique to minimize recall bias and improve accuracy. Participants will be asked to list all fruits and vegetables consumed from waking to bedtime, including portion sizes estimated using food models, household measures, and photographs. Special emphasis will also be placed on capturing the type, source, preparation method, and the quantity of fruits and vegetables consumed.
- The list on the QFFQ will include all fruits and vegetables targeted in this study and the commonly consumed ones from the 24HR. For each item, participants will report the Frequency of consumption over the past month using seven response categories: never, 1-3 times per month, once per week, 2-4 times per week, 5-6 times per week, once per day, and 2 or more times per day and the usual portion size consumed per occasion, using local household measures and food photographs for estimates [36][37].

### Human Health Risk Assessment

#### Estimated Daily Intake (EDI)

Average daily consumption of each and vegetable obtained from the 24-hour recalls and QFFQ will be combined with measured PTTE concentrations to estimate individual dietary exposure. The average concentration of PTTEs in vegetables and fruits multiplied by the food intake rate of fruit and vegetables (g/person/day) will be employed to calculate daily intakes (EDIs) of PTTEs (mg/day) [38]. One common method that people are exposed to PTTEs is through eating contaminated fruits and vegetables daily. The following equation will be employed to calculate the EDI for the PTTEs from the fruits and vegetables in the study [39].

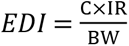

Where:

EDI = Estimated Daily Intake (mg/kg/day)
C = Concentration of PTTEs in fruits and vegetables (mg/kg, dry weight) IR = Ingestion rate of fruits and vegetables (kg/person/day)
BW = Average adult body weight (70kg) [40]

#### Target Hazard Quotient (THQ)

The following equation will be employed to determine the THQ, a ratio that defines the danger of non-carcinogenic repercussions for each PTTE through fruits and vegetables ingestion. The non-cancer hazards are represented by THQ, the exposure frequency (365 days/year) by EF, the exposure length (70years) by ED, the average time for non-carcinogens (AT=365×ED) by AT, and the reference dose for oral ingestion by RfD. In terms of non-carcinogenic hazards, THQ levels below 1 are typically regarded as tolerable. Noncarcinogenic effects are possible, nevertheless, if the value is greater than 1, and this likelihood rises with the value [41].

The oral reference doses (RfD) for each PTTE are presented below [40]

| Potentially Toxic Trace Element | RfD (mg/kg/day) |
| --- | --- |
| Arsenic, As | 0.00032 |
| Cadmium, Cd | 0.001 |
| Lead, Pb | 0.004 |
| Zinc, Zn | 0.30 |

### Hazard Quotient (THQ/HQ) and Hazard Index (HI)

#### Target Hazard Quotient (THQ)

The Target Hazard Quotient (THQ) is the ratio of the determined dose of a contaminant to the oral reference dose. The equation below will be employed to estimate the noncarcinogenic risk of PTTEs through consumption of contaminated fruits and vegetables [41][42].

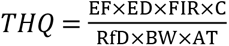

Where THQ = Target Hazard Quotient (Unitless)

ED = Exposure Duration (70 years)
EF = Exposure Frequency (365 days/year)
FIR = Food Ingestion rate (kg/person/day), equivalent to IR
C = Concentration of PTTEs (mg/kg)
RfD = Oral reference dose (mg/kg/day)
BW = Average body weight (70kg)
AT = Average time for non-carcinogens (EF×ED= 25,550 days)

For the study, exposure will be assumed to be daily, therefore EF×ED=AT. The model simplifies to the widely used equation [38]

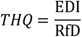

A THQ value < 1 indicates no appreciable non-carcinogenic risk, while THQ > 1 indicates a potential health risk.

#### Hazard Index (HI)

The Hazard Index (HI) will be employed to assess the cumulative noncarcinogenic risk to human health from exposure to multiple PTTEs in the fruits and vegetables. The population may have a negative health implication if the HI value is greater than 1. The formula below will be employed to determine the HI Value [43].

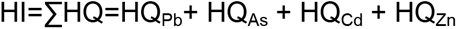

Where each fruit and vegetable species has the specific PTTEs Pb, Cd, As and Zn.

#### Target Carcinogenic Risk (TCR)

The lifetime Target Carcinogenic Risk (TCR) will be calculated for the carcinogenic elements to estimate the risk of cancer over a lifetime due to the exposure through the consumption of fruits and vegetables. The lifetime target TCR can be calculated as

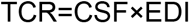

Where:

TCR= Target Carcinogenic Risk, unitless
CSF= Carcinogenic Slope Factor (mg/kg/day)^-1^
EDI= Estimated Daily Intake (mg/kg/day)

Where, TCR represents the probability of an individual developing cancer over a lifetime due to exposure to a carcinogenic element. The slope factor of, As, Cd, and Pb are 1.5,6.1 and 8.5 ×10^-3^ (mg/kg/day)^-1^, respectively. The target carcinogenic risk greater than 1.0 x 10^-4^ to be considered intolerable. Zn does not have an oral CSF so it will be excluded from the TCR calculations but will be assessed for non-carcinogenic risk using THQ.

### Statistical Analysis

Data will be analysed using IBM SPSS version 29 and the normality of the PTTEs concentrations in the fruits and vegetables will be assessed using a Shapiro-Wilk test. Results will be presented as the mean ± standard deviation if normally distributed or median (interquartile range) if not normally distributed.

If results will be normally distributed, one-way ANOVA will be used to determine differences in the concentrations of each PTTE separately across different food types and between food groups followed by Tukey post-hoc test for multiple comparisons. If data are not normally distributed, the Kruskal-Walli’s test followed by Mann-Whitney U test with Bonferroni correction will be used. Where necessary, multivariate approaches will be considered to explore overall patterns of contamination.

### Dietary Analysis

Data from the administered 24-hour recalls and the QFFQS will be recorded and analysed using Food Finder 3 Software linked to the South African Food Composition Database [44]. All foods and beverages reported in the 24-hour recalls will be coded and matched to items in SAFOODS. For some fruits and vegetables that are indigenous and consumed in Thulamela Municipality and are not listed SAFOODS, nutrient values will be imputed from USDA food Data Central of published South African studies. Mean daily intake of energy, macronutrients and micronutrients contributed by fruits and vegetables will be calculated from the 24-hour recall day.

The usual daily intake from the QFFQ of each fruit and vegetable will be estimated by converting the reported frequency and the portion sizes into grams per day using the formula: Frequency per day × Usual portion size (g). Total fruit intake, total vegetable intake, and combined fruits + vegetable intake will be summed for each participant. Total intake will be compared to the WHO recommendations [45]. Data will be exported to IBM SPSS Statistics version 29 for consistency with the Food Finder 3 output-format.

Descriptive statistics will be used to summarize dietary intake data. After the Shapiro-Wilk test is used to determine if the data is normally distributed, means and standard deviations will be supplied, and medians and interquartile ranges will be reported for non-normally distributed. Each participant’s total fruit, total vegetable, and combination fruit and vegetable intake (g/day) will be determined. The proportion of participants who consume at least 400 grams of fruits and vegetables per day, as recommended by WHO, will be determined. Statistical significance will be established at p<0.05 and associations will be evaluated using Pearson’s correlation for normally distributed data or Spearman’s rank correlation for non-normally distributed, as appropriate.

### Ethical Considerations

Ethical clearance will be obtained from the University of Venda, Faculty of Health Sciences Research Ethics Committee and Gatekeeper permission will be sought from Thulamela Municipality to conduct research within the municipality. Written permission will be sought from the management of the selected supermarkets in Thulamela Municipality. Informed consent will be obtained from the participants before administering 24-hour recalls and QFFQS, which will be available in English and Tshivenda. The consent process will include detailed information and the purpose of the study, the procedures involved and any risks or benefits. Participants will be assured of the confidentiality and voluntary nature of their participation, with the right to withdraw at any time without consequences. No biological samples will be collected from participants.

All hardcopy data will be stored in a locked filing cabinet in the supervisor’s office at the University of Venda. Electronic data will be password-protected and will only be accessible by the researcher and the supervisor. Data will be kept for 5 years after publication

The study will adhere to the ethical principles outlined in the Declaration of Helsinki and the South African National Health Act no. 61 of 2003 [46][47].

## Data Availability

All relevant data are within the manuscript and its Supporting Information files.

## Plan for dissemination of results

Study findings will be disseminated through publication in an open-access, peer-reviewed journal to ensure wide accessibility. Key results will be presented at provincial and national public health and nutrition conferences. A summary report will be shared with the Thulamela Local Municipality, Vhembe District Health Office, Limpopo Department of Health, and Department of Nutrition to inform food safety and nutrition policies and the study results will also be communicated back to participating communities in Thulamela through community feedback meetings and information leaflets in English and Tshivenda.

## Acknowledgements

The authors would like to acknowledge the Thulamela Local Municipality, Vhembe District Health Office, and Department of Nutrition for their support. We thank the community members in Thulamela for their anticipated participation. Funding for this study will be sought from relevant research funding bodies. Ethics approval will be sought from the University of Venda Human and Social Sciences Research Ethics Committee prior to commencement of the study.

